# COVID-19 Vaccine Effectiveness against Symptomatic and Asymptomatic SARS-CoV-2 Infections with the Delta Variant among a Cohort of Children Aged ≥ 12 Years and Adults in Utah

**DOI:** 10.1101/2022.10.13.22281012

**Authors:** Sarita Mohanty, Fatimah S. Dawood, Joseph B. Stanford, Jazmin Duque, Melissa S. Stockwell, Vic Veguilla, Christina A. Porucznik

## Abstract

We conducted weekly surveillance for SARS-CoV-2 infection among a sample of households with ≥1 child aged 0-17 years from selected Utah counties. A Cox proportional hazards model approach was used to calculate infection hazard rate and vaccine effectiveness. Findings show that the recommended primary series of COVID-19 vaccine was effective against circulating variants during a Delta-predominant wave in Utah.

## Introduction

Despite evidence that COVID-19 vaccines protect against SARS-CoV-2 infections, hospitalizations, and deaths,^1^ COVID-19 vaccine uptake has remained low in some segments of the U.S. population. Concern about whether COVID-19 vaccines are effective is one of several barriers to vaccination among individuals who are vaccine hesitant.^2^ Even as the recommended COVID-19 vaccine series evolves to include booster doses, COVID-19 vaccine effectiveness (VE) estimates from diverse communities and time periods remain important to build an evidence base to inform individual choices about vaccination. Using data from a household cohort under intensive surveillance for asymptomatic and symptomatic SARS-CoV-2 infection, we estimate COVID-19 VE against infection among participants aged ≥12 years during a SARS-CoV-2 Delta variant-predominant wave in Utah.

## Materials and Methods

The Coronavirus Household Evaluation and Respiratory Testing (C-HEART) study conducted surveillance for SARS-CoV-2 infection among a convenience sample of households with ≥1 child aged 0-17 years from selected Utah counties (Salt Lake, Weber, Davis, Box Elder, Cache, Tooele, Wasatch, Summit, Utah, and Iron) during September 2020–August 2021.^3^ The C-HEART surveillance period included a Delta variant-predominant circulation wave (Figure 1).^4^ Every week, participants self-collected (or adult caregivers collected from children) mid-turbinate nasal swabs (MTS) for reverse transcription polymerase chain-reaction testing for SARS-CoV-2 and lineage typing and responded to symptom questionnaires. COVID-19 vaccination status was ascertained from participants’ self-report and/or verification with vaccine cards or the Utah State Immunization Information System. Participants self-collected additional MTS with the onset of COVID-19-like illness symptoms (Supplemental Methods). The study protocol was reviewed and approved by the University of Utah Institutional Review Board (IRB) which served as the central IRB for all study collaborators.

**Figure 1:**
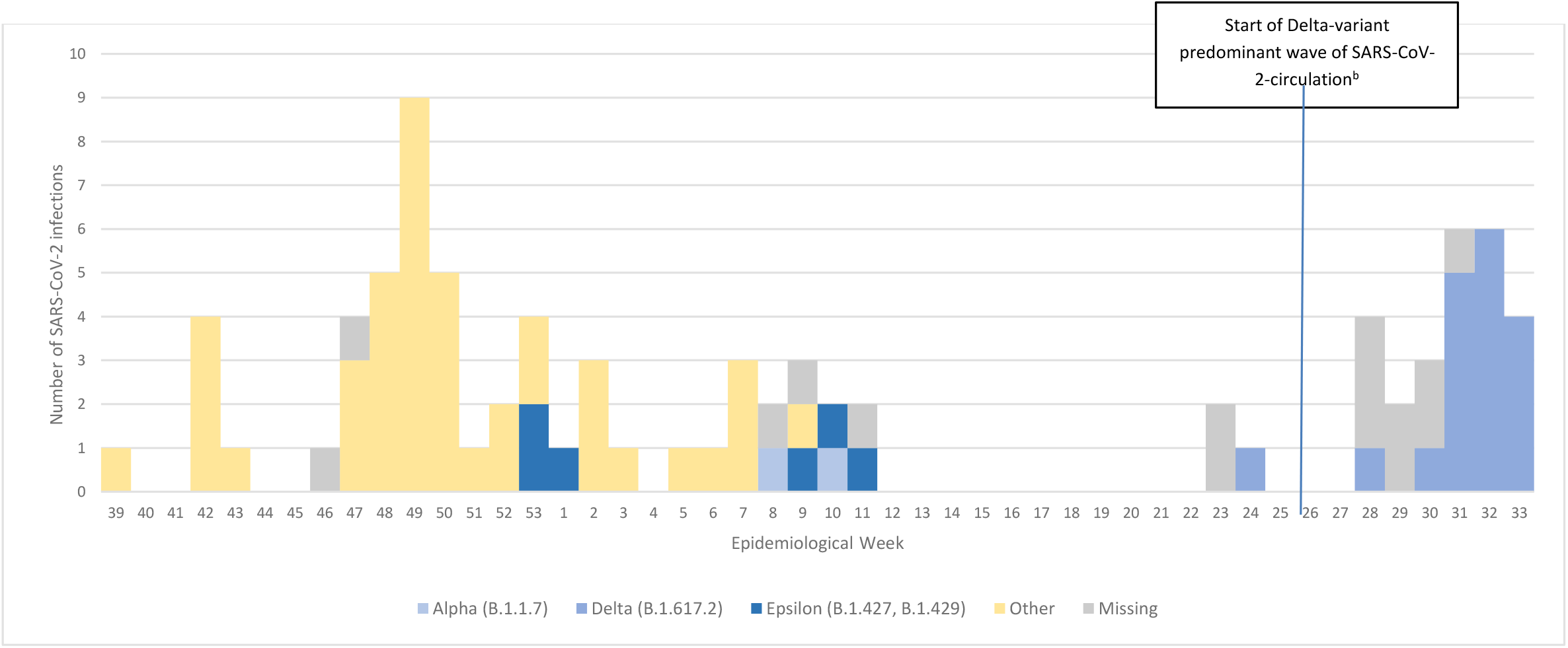
SARS-CoV-2 infections by variant lineage among cohort participants of all ages^a^, August 2020–February 2022, n=84 infections. ^a^ Figure shows infections among all age groups but there were 13 infections among individuals ≥12 years during the Delta-predominant wave ^b^ The Delta-predominant wave of SARS-CoV-2 circulation in Utah started on June 27, 2020 (epidemiological week 26)

This analysis was restricted to participants aged ≥12 years as of their enrollment into the cohort because the FDA approved Emergency Use Authoriazations for COVID-19 vaccines for persons aged ≥16 years in December, 2020 and for children 12-15 years in May, 2021. The analysis period spans June 13, 2021 (the start of the Delta predominant wave indicated by the first Delta infection case in the cohort) through August 21, 2021, when cohort surveillance ended. Participants who previously had RT-PCR-confirmed SARS-CoV-2 infection prior to June 13, 2021 were excluded from the analysis. Demographic characterstics of vaccinated versus unvaccinated participants were compared using Chi Square and Fisher’s exact test. Cox proportional hazards models with robust standard errors to account for household clustering were used to calculate infection hazard rate (HR) ratios during unvaccinated versus fully vaccinated person-time, after adjusting for age group and demographic characteristics that were available for both age groups and differed among vaccinated versus unvaccinated individuals in bivariate analysis at a p-value threshold of <0.1 (highest educational level of adults in the household and having at least one child <12 years in the household) (Supplemental Methods). Models were structured using epidemiologic weeks as the time scale. Vaccination status was treated as a time-varying covariate. Participants were considered fully vaccinated at ≥14 days after recieving all recommended primary COVID-19 vaccine doses. Weeks between the first vaccine dose and full vaccination status were excluded. VE was calculated as (1-HR)*100.

## Results

As of June 13, 2021, 384 participants aged ≥12 years were under surveillance, including 70 children aged 12-15 years and 314 persons aged ≥16 years (age range 16-70) (Table 1). Among the 384 participants, 337 (88%) were non-Hispanic White and 361 (94%) were fully vaccinated against COVID-19 before or during the analytic period. Ninety-six percent (341/357) of fully vaccinated participants lived in households with at least one adult with a college education, compared to 85% (17/23) of unvaccinated participants (p=0.001)

**Table 1.** Baseline characteristics^a^ of C-HEART participants aged ≥12 years under SARS-CoV-2 surveillance, June 13, 2021 to August 21, 2021by COVID-19 vaccination status, N=384.

|  | All participants |  | Vaccinated<sup>c</sup> |  | Unvaccinated<sup>c</sup> |  | p-value<sup>g</sup> |
| --- | --- | --- | --- | --- | --- | --- | --- |
|  | n=384 |  | n=361 |  | n=23 |  |  |
|  | n | col % | n | col % |  | col % |  |
| **All individuals** |  |  |  |  |  |  |  |
| Age group |  |  |  |  |  |  |  |
| 12-15 years | 70 | 18% | 66 | 18% | 4 | 17% | 1.000 |
| $\geq 16$ years | 314 | 82% | 295 | 82% | 19 | 83% | |
| Gender |  |  | 0 |  |  |  |  |
| Male | 186 | 48% | 174 | 48% | 12 | 52% | 0.712 |
| Female | 198 | 52% | 187 | 52% | 11 | 48% |  |
| Non-binary/third gender | 0 | 0% | 0 | 0% | 0 | 0% |  |
| Race/ethnicity<sup>b</sup> |  |  |  |  |  |  |  |
| Black, Non-Hispanic | 2/383 | 1% | 1/360 | 0% | 1/23 | 4% | 0.171 |
| Hispanic | 22/383 | 6% | 22/360 | 6% | 0/23 | 0% |  |
| Other, Non-Hispanic | 22/383 | 6% | 21/360 | 6% | 1/23 | 4% |  |
| White, Non-Hispanic | 337/383 | 88% | 316/360 | 88% | 21/23 | 91% |  |
| $\geq 1$ medical condition<sup>c</sup> | 166 | 43% | 157 | 43% | 9 | 39% | 0.682 |
| Asthma | 72 | 19% | 71 | 20% | 1 | 4% |  |
| Other medical conditions besides asthma | 94 | 24% | 86 | 24% | 8 | 35% |  |
| Weeks from fully vaccination status to Delta wave start |  |  | 9.6 | [5.4 - 15.4] |  |  |  |
| 12-15 years |  |  | 1.1 | [0.9 - 1.4] |  |  |  |
| $\geq 16$ years | | | 10.6 | [7.3 - 16.4] | | | |
| Highest educational level attained by adults aged $\geq 18$ years in the household<sup>d</sup> | | | | | | | |
| Less than high school graduate | 0/380 | 0% | 0/357 | 0% | 0/23 | 0% | 0.001 |
| High school graduate | 1/380 | 0% | 1/357 | 0% | 0/23 | 0% |  |
| Some college or technical school | 21/380 | 6% | 15/357 | 4% | 6/23 | 26% |  |
| College graduate | 358/380 | 94% | 341/357 | 96% | 17/23 | 74% |  |
| Lives in the same household with $\geq 1$ child under the age of 12 | 307 | 80% | 285 | 79% | 22 | 96% | 0.059 |
| **Adults (aged $\geq 18$ years, n=298)** |  |  |  |  |  |  |  |
| Employment and telework status |  |  |  |  |  |  |  |
| Employed, teleworking | 113/292 | 39% | 111/276 | 40% | 2/16 | 13% | 0.038 |
| Employed, not teleworking | 129/292 | 44% | 120/276 | 43% | 9/16 | 56% |  |
| Not employed | 50/292 | 17% | 45/276 | 16% | 5/16 | 31% |  |
| **Children (aged 12-17 years, n=86)** |  |  |  |  |  |  |  |
| School outside the home during cohort participation | 80/86 | 93% | 74/80 | 93% | 6/6 | 100% | 0.487 |

Denominators indicate individuals with available data for each variable if different from total number of individuals. 1
person had missing race data; 4 had missing highest education level attained by adults aged >18 years in the household data; and 6 adults aged $\geq 18$ years had missing employment and telework status data
<sup>a</sup> All characteristics were ascertained at enrollment only unless otherwise specified.
<sup>b</sup> Other, non-Hispanic includes participants who self-identified as non-Hispanic and Asian, Native Hawaiian/Pacific Islander, or multiple races.
<sup>c</sup> Based on questionnaires that asked about the following medical conditions: asthma, chronic lung disease other than asthma, chronic metabolic diseases such as diabetes mellitus types 1 and 2 and thyroid disease, blood disorders such as thalassemia and sickle cell disease, hypertension, cardiovascular disease, bladder/kidney disease, liver disease, immunocompromising conditions, neurologic/neuromuscular disease and rheumatologic conditions.
<sup>d</sup> Only adults aged $\geq 18$ years were asked about their highest educational level at study enrollment.
<sup>e</sup> Based on vaccine receipt at the end of the study period on August 21, 2021. Participants were defined as vaccinated if they either self-reported receiving all recommended primary doses of COVID-19 vaccine and/or were documented as having received vaccine based on participant-submitted COVID-19 vaccine cards or Utah State Vaccine Registry Data.
<sup>f</sup> Based on weekly questionnaires about school attendance outside the home collected throughout the study period.
<sup>g</sup> p-values were calculated based on a Chi-square test unless a cell frequency < 5, in which case a Fisher's exact test was used

The 384 participants were followed for 3042 person-weeks (median per participant, 8.6 weeks, IQR 7.3-9.6) during the analytic period, including 161 unvaccinated weeks and 2880 fully vaccinated weeks. Fourteen SARS-CoV-2 infections were identified, including 11 among fully vaccinated and 3 among unvaccinated participants. Of the fourteen infections, two were among children aged 12-15 years (one vaccinated and one unvaccinated). Of the 11 infections among fully-vaccinated participants, the median time from full vaccination status to infection was 9.6 weeks (IQR 5.4-15.4). Six of eleven infections among fully-vaccinated participants were asymptomatic (versus 0 of 3 infections among unvaccinated participants, p=0.26). Adjusted VE against any SARS-CoV-2 infection among all participants aged ≥12 years was 85% (95% CI 46-96).

## Conclusion

During a Delta variant-predominant wave of SARS-CoV-2 circulation in Utah, a primary series of a COVID-19 vaccine was effective at preventing SARS-CoV-2 infection (both asymptomatic and symptomatic) in persons aged ≥12 years. There was a trend towards a larger proportion of SARS-CoV-2 infections in fully vaccinated individuals being asymptomatic, suggesting that COVID-19 vaccination may attenuate symptom status; however, infection numbers were small, precluding VE estimates by symptom status. Our findings are consistent with other published estimates of the effectiveness of a COVID-19 vaccine primary series against Delta variant infections (range 78-84%).^1^ Findings also suggest that COVID-19 vaccines may be effective against asymptomatic and symptomatic infections, which is critical to reducing community transmission.

Several limitations should be considered when interpreting analysis findings. First, as previously shown,^3^ the distribution of selected demographic characteristics (race, educational level, household education) differs between the cohort and the general U.S. population, so findings may not be fully generalizable to all U.S. communities. However, our estimates of VE are consistent with estimates from studies in a variety of populations. Second, a small sample size also precluded us from assessing VE stratified by symptom status, age group, or time since vaccination and also resulted in wide confidence intervals. Third, findings from this analysis are specific to VE against the Delta variant and are not generalizable to VE against more recent emerging variants. However, our findings provide evidence that the recommended primary series of COVID-19 vaccine was effective against circulating variants during the Delta-predominant wave study period.

## Data Availability

All data produced in the present study are available upon reasonable request to the authors

## Declarations of Interest

none

## Role of the funding source

CDC funded this study. CDC-affiliated authors were involved in study design, data collection, analysis and interpretation, report writing, and the decision to submit the paper for publication. The corresponding author had full access to all data used in the analysis and had final responsibility for the decision to submit for publication.

## Acknowledgements

The authors wish to acknowledge the families that participated in the C-HEART cohort; the REDCap data platform; Jacob Anderson, Kayley Cheminant, Halle Fiagle, Kristina Gale, Kathryn Graham, and Emily Hacker from the University of Utah; Priyam Thind, Maria Castro, Franklin Sosa, Luis Alba, Liqun Wang, Chelsea Wynn, John Paul Harris, Laura Staeheli, Angela Cameron, and Ogooluwa Fayemiwo from Columbia University; and Danielle Rentz Hunt, Zuha Jeddy, Kim Altunkaynak, Utsav Kattel, Parler Malek, Hannah Michelle Brower, Tana Brummerand Brandon Poe from Abt Associates. Study data were collected and managed using REDCap electronic data capture tools hosted at Vanderbilt University Medical Center with grant support UL1 TR000445 from NCATS/NIH. REDCap (Research Electronic Data Capture) is a secure, web-based software platform designed to support data capture for research studies, providing 1) an intuitive interface for validated data capture; 2) audit trails for tracking data manipulation and export procedures; 3) automated export procedures for seamless data downloads to common statistical packages; and 4) procedures for data integration and interoperability with external sources.

## Funding

This study was funded by the US Centers for Disease Control and Prevention through Contract # 75D30120C08150 with Abt Associates.

## Notes

### Competing Interest Statement

The authors have declared no competing interest.

### Author Declarations

The study protocol was reviewed and approved by the University of Utah Institutional Review Board (IRB) which served as the central IRB for all study collaborators

### Summary of Updates

Title has been edited to make the study population clearer

## References

1. Pormohammad A, Zarei M, Ghorbani S, Mohammadi M, Aghayari Sheikh Neshin S, Khatami A, et al. (2021). Effectiveness of COVID-19 Vaccines against Delta (B.1.617.2) Variant: A Systematic Review and Meta-Analysis of Clinical Studies. Vaccines, 10(1), 23. MDPI AG. Retrieved from http://dx.doi.org/10.3390/vaccines10010023

2. Nguyen KH, Srivastav A, Razzaghi H, Williams W, Lindley M, Jorgensen C, et al. COVID-19 Vaccination Intent, Perceptions, and Reasons for Not Vaccinating Among Groups Prioritized for Early Vaccination — United States, September and December 2020. MMWR Morb Mortal Wkly Rep 2021;70:217–222. DOI: http://dx.doi.org/10.15585/mmwr.mm7006e3

3. Dawood FS, Porucznik CA, Veguilla V, Stanford JB, Duque J, Rolfes MA, et al. Incidence Rates, Household Infection Risk, and Clinical Characteristics of SARS-CoV-2 Infection Among Children and Adults in Utah and New York City, New York. JAMA Pediatr. 2022;176(1):59–67. DOI: http://dx.doi.org/10.1001/jamapediatrics.2021.4217

4. Centers for Disease Control and Prevention. COVID Data Tracker. Atlanta, GA: US Department of Health and Human Services, CDC; 2022, March 11. https://covid.cdc.gov/covid-data-tracker

5. Paden CR, Tao Y, Queen K, Zhang J, Li Y, Uehara A … Tong S (2020). Rapid, Sensitive, Full-Genome Sequencing of Severe Acute Respiratory Syndrome Coronavirus 2. Emerging infectious diseases, 26(10), 2401–2405. DOI: http://dx.doi.org/10.3201/eid2610.201800

6. 2019. BBMap. SourceForge. https://sourceforge.net/projects/bbmap

7. Aksamentov I, Roemer C, Hodcroft EB, and Neher RA, (2021). Nextclade: clade assignment, mutation calling and quality control for viral genomes. Journal of Open Source Software, 6(67), 3773, http://dx.doi.org/10.21105/joss.03773

8. O’Toole A, Scher E, Underwood A, Jackson B, Hill V, McCrone JT, et al., Assignment of epidemiological lineages in an emerging pandemic using the pangolin tool, Virus Evolution, Volume 7, Issue 2, September 2021, veab064, http://dx.doi.org/10.1093/ve/veab064

