## Supplemental Methods for "COVID-19 Vaccine Effectiveness against Symptomatic and Asymptomatic SARS-CoV-2 Infections with the Delta Variant among a Cohort of Children Aged ≥ 12 Years and Adults in Utah"

*Surveillance for Symptoms associated with SARS-CoV-2 Infection*

Cohort participants were contacted by text message or email every week to ascertain whether they had COVID-19-like illness (CLI) symptoms or any other illness symptoms. CLI was defined as ≥1 symptoms of fever or feverishness, cough, shortness of breath, sore throat, diarrhea, muscle aches, chills, or change in taste or smell. Participants who reported illness symptoms during the initial weekly surveillance screening question completed an additional questionnaire about specific symptoms including CLI symptoms plus joint pain, nausea, vomiting, abdominal pain, headache, eye redness, nasal congestion/runny nose, rashes including skin changes on fingers or toes or other rashes, chest pain, abnormal fatigue, and increased fussiness/inconsolable crying in children aged <2 years. For this analysis, SARS-CoV-2 infections were classified as asymptomatic if individuals did not report any symptoms associated with the infection during the 7 days preceding their first positive mid-turbinate nasal swab sample through the last week in which they had a positive swab sample.

*COVID-19 Vaccination Ascertainment*

In early 2021, after COVID-19 vaccines received FDA emergency use authorization (EUA) in the United States, the weekly surveillance questionnaire for participants aged ≥16 years included additional questions every fourth week to collect information about COVID-19 vaccine receipt, vaccine type, and date of receipt. The additional questions were also asked of participants aged 12-15 years once the FDA EUA was expanded to include approval of COVID-19 vaccines for this age group. Participants who reported receiving COVID-19 vaccine were asked to upload their COVID-19 vaccination card for study staff to verify self-reported responses. At the end of the study period, the study team also collected COVID-19 vaccination information for all vaccine-eligible participants from the Utah State Registry.

*SARS-CoV-2 Sequencing*

Participants with SARS-CoV-2-positive samples with Cycle Threshold values <30 had samples processed for whole genome sequencing at the Centers for Disease Control and Prevention by previously published methods ^5^ or using the IDT xGen SARS-CoV-2 library prep kit (Integrated DNA Technologies, Inc., Coralville, IA). Libraries were sequenced using 2x150 base pair Illumina Chemistry on a MiSeq or NovaSeq instrument (Illumina Inc., San Diego, CA). Demultiplexed data were down-sampled to 1 million reads per sample, primers were trimmed with BBDuk ^6^ and a single consensus genome for each sample was generated with IRMA v1.0.2 using the default CoV configuration. Clade assignments were made using Nextclade version 1.13.2 ^7^ and Pangolin version 3.1.20. ^8^
